# Did COVID-19 or COVID-19 vaccines influence the patterns of Dengue in 2021: An exploratory analysis of two observational studies from North India

**DOI:** 10.1101/2023.01.09.23284366

**Authors:** Upinder Kaur, Parth Jethwani, Shraddha Mishra, Amol Dehade, Ashish Kumar Yadav, Sasanka Chakrabarti, Sankha Shubhra Chakrabarti

**Affiliations:** Department of Pharmacology, Institute of Medical Sciences, Banaras Hindu University, Varanasi, Uttar Pradesh, India; Department of Endocrinology and Metabolism, All India Institute of Medical Sciences, Jodhpur, Rajasthan, India; Center for Biostatistics, Institute of Medical Sciences, Banaras Hindu University, Varanasi, UP, India; Department of Biochemistry and Central Research Cell, Maharishi Markandeshwar (deemed to be) University, Mullana, Haryana, India; Department of Geriatric Medicine, Institute of Medical Sciences, Banaras Hindu University, Varanasi, Uttar Pradesh, India

**Keywords:** Dengue, Non-COVID infections, long COVID, Vaccine after COVID, COVID-19 Vaccines

## Abstract

**Background:** Dengue which is endemic in India and has been occurring for decades apparently witnessed a rise in disease burden in 2021 in specific regions of the nation. We aim to explore less studied risk factors of Dengue occurrence and severity in the post-COVID-19 and post-COVID-19 vaccination era.

**Methods:** This was an exploratory analysis involving participants from two prior observational studies conducted during the period of Feb 2021-April 2022 in a tertiary hospital in North India. Healthcare workers constituted the majority of study participants. Individuals were stratified into five groups based on COVID-19 infection and timing of vaccination: CovidNoVaccine (CNV), VaccineNoCOVID (VNC), CovidAfterVaccine (CAV), VaccineAfterCOVID (VAC) and NoVaccineNoCovid (NVNC) groups. The occurrence of lab-confirmed Dengue and severe forms of Dengue were the main outcomes of interest. We tried to predict determinants of Dengue occurrence and severity with a particular focus on COVID-19 history and vaccination status.

**Results:** A total of 1520 vaccinated individuals and 181 unvaccinated individuals were included. Of these 1701 participants, symptomatic Dengue occurred in 133 (7.8%) and was of ‘severe’ category in 42 (31.6%). Individuals with a history of COVID-19 in 2020 had 2 times higher odds of developing symptomatic Dengue. The VAC group had 3.6, 2- and 1.9 times higher odds of developing Dengue than the NVNC, VNC, and CAV groups. The severity of dengue was not affected by COVID-19 or COVID-19 vaccination.

**Conclusions:** COVID-19 may enhance the risk of developing symptomatic dengue. Future research dealing with long COVID should explore the propensity of COVID-19 victims towards symptomatic forms of other viral illnesses. Individuals receiving the COVID-19 vaccine after recovering from COVID-19 particularly seem to be at greater risk of symptomatic dengue and need long-term watchfulness. Possible mechanisms, such as antibody-mediated enhancement or T-cell dysfunction, should be investigated in COVID-19-recovered and vaccinated individuals. Further large-scale, multicentric, robust studies with a better enrolment of unvaccinated people will help understand the interplay of factors involved in COVID-19 and Dengue.

## 1. Introduction

As of 25th Nov 2022, COVID-19 affected more than 0.6 billion people globally and claimed above 6 million lives (1). In the fight against the virus, COVID-19 vaccination programs were started across the world in late 2020. In India, the mass rollout of COVID-19 vaccines was initiated in January 2021 primarily employing the ChAdOx1-nCoV-19 vaccine (COVISHIELD, Serum Institute of India under license from Oxford-AstraZeneca), and the inactivated SARS-CoV-2 vaccine, BBV152 (COVAXIN, Bharat Biotech). The country however was hit by SARS-CoV-2 during the 2^nd^ wave of the pandemic which started in the mid of March 2021 and peaked in the first week of May 2021 with daily confirmed cases of COVID-19 reaching as high as 0.38 million. In this regard, we previously reported a high rate (25-40%) of breakthrough infections in the ChAdOx1-nCoV-19 vaccinated cohort primarily composed of healthcare workers of our institute (2). The virulent delta strain of SARS-CoV-2 was considered responsible for the devastating second wave of the COVID-19 pandemic (3). In the following months, from July to September 2021, coinciding with the seasonal trends, a surge of Dengue cases was observed in the country with the reported burden and deaths being more than four and six times respectively compared to the year 2020 (4). In Uttar Pradesh, the numbers were even more substantial with the reported number of cases more than eight times compared to 2020 (4). The authors also witnessed a similar high rate of infections and hospital admissions because of Dengue in health care workers, a pattern that was not observed over the last five years. The reasons and determinants of these disturbing trends of occurrence and severity of Dengue have not been adequately studied. The possible modulation of patterns of Dengue by COVID-19 and COVID-19 vaccination has been unexplored. The present study aims to determine the risk factors of Dengue occurrence and severity in the post COVID-19 and-post COVID-19 vaccination era. Particularly, we have tried to explore the effect of COVID-19 and COVID-19 vaccination on the Dengue occurrence and severity.

## 2. Materials and methods

### 2.1. Study design and participants

The present study is an exploratory analysis and involved participants from two studies conducted in the Sir Sunder Lal Hospital which is a large tertiary care referral center in northern India. The first study was a prospective observational safety study of individuals vaccinated with ChAdOx1-nCoV-19 and was conducted from the period of February 2021 to April 2022. The safety of ChAdOx1-nCoV-19 and occurrence of COVID-19 following vaccination were the main outcomes of the study and preliminary findings have been published by us last year (2,5). Being the priority vaccine recipients in the first phase of COVID-19 vaccination, healthcare workers and the elderly constituted the study population. In view of the observed variations in patterns of Dengue in the institute, Dengue was incorporated as a separate entity in the list of adverse events of special interest (AESIs) in the final follow-up of this one year-long safety study (6). The severity of Dengue as an AESI was defined in accordance with the FDA rating of adverse events following immunization. Any form of Dengue that required intravenous fluid therapy at home (FDA grade ‘severe’) or resulted in hospitalization (FDA grade ‘serious’) with or without the need of platelet transfusion, was clubbed under ‘severe’ Dengue. In order to make meaningful comparisons of the vaccinated cohort from this study with an unvaccinated cohort with similar demographic and occupational risk factors, we drew unvaccinated participants from a second study whose findings have also been published by us previously. This second study was based on a retrospective cohort design with the primary objective of determining COVID-19 vaccine effectiveness and thus enrolled both vaccinated and unvaccinated health care workers of the institute (7). This study was conducted from July 2021 to December 2021. The unvaccinated individuals from this study were contacted again during the months of October-November 2022 to obtain the Dengue related information. Vaccination status was again confirmed from these individuals as it was possible that they might have received the COVID-19 vaccines in the intervening period. For the present analysis, unvaccinated individuals were defined as those who had not taken any dose of COVID-19 vaccine till the major curve of Dengue cases touched the baseline (till 30^th^ Sept 2021 based on data collected from the study participants), mentioned hereafter as the ‘end date’. All those who had taken at least one dose of COVID-19 vaccine before 1^st^ July 2021 which coincided with the start of the Dengue epidemic (mentioned hereafter as the ‘start date’) were clubbed together in the vaccinated arm. To prevent adulteration by cases who received vaccine during the Dengue epidemic period (1^st^ July 2021 to 30^th^ September 2021), individuals receiving their first dose of vaccine during this period were excluded from the present analysis.

To understand the cumulative effect of COVID-19 occurring at any time, on Dengue occurrence and severity, and to explore the effect of timing of COVID-19 vaccine with respect to COVID-19 illness on Dengue occurrence and severity, participants were categorized into five groups as defined below:

### Categorization A

1. Covid-No Vaccine (CNV) group: Individuals who had a history of COVID-19 before the ‘start date’ but were unvaccinated till the ‘end date’
2. Vaccine-No COVID (VNC) group: Individuals who had received at least one dose of COVID-19 vaccine before the ‘start date’ but had no history of COVID-19 till the ‘start date’
3. Covid After Vaccine (CAV) group: Individuals who after receiving COVID-19 vaccine, developed COVID-19 at any time but before the ‘start date’
4. Vaccine After COVID (VAC): Individuals who had a history of COVID-19 in the year 2020 and then received COVID-19 vaccine in the year 2021 at any time but before the ‘start date’
5. No Vaccine No Covid (NVNC) group: Individuals who had no history of COVID-19 till the ‘start date’ and were unvaccinated till the ‘end date’.

### Categorization B

As mentioned above, the Institute was hit by the 2^nd^ wave of COVID-19 pandemic in the months of March -May 2021. Also, as per the revised recommendations by the authorities on COVID-19 vaccination, the interval between two doses of COVID-19 vaccine was extended to 84 days. Consequently, some participants received their 2^nd^ dose of COVID-19 vaccine post recovery from COVID-19 which they had in the year 2021. The individuals who had received their 2^nd^ dose of COVID-19 vaccine after COVID-19 of the year 2021, but before the development of Dengue, were shifted from the CAV group of Categorization A to the VAC group under Categorization B and analysed separately.

### 2.2. Ethical concerns

Ethical permission was obtained from the Institute Ethics Committee and informed consent was taken from the study participants.

### 2.3. Outcomes

Rate of occurrence of lab confirmed Dengue and rate of severe forms of Dengue were the main outcomes studied in the present analysis. In addition, we predicted the determinants of Dengue occurrence and severity with particular focus on COVID-19 history and vaccination status.

## 3. Results

Figure 1 shows the flowchart of study participants. Out of 1650 participants from study 1 who were vaccinated with the ChAdOx1-nCoV-19 vaccine, 1520 could be contacted for Dengue analysis. As mentioned earlier, healthcare workers of the institute dominated the group. From study 2 conducted in the same institute, out of 216 unvaccinated healthcare workers, 181 were included for the present analysis.

**Figure 1.**
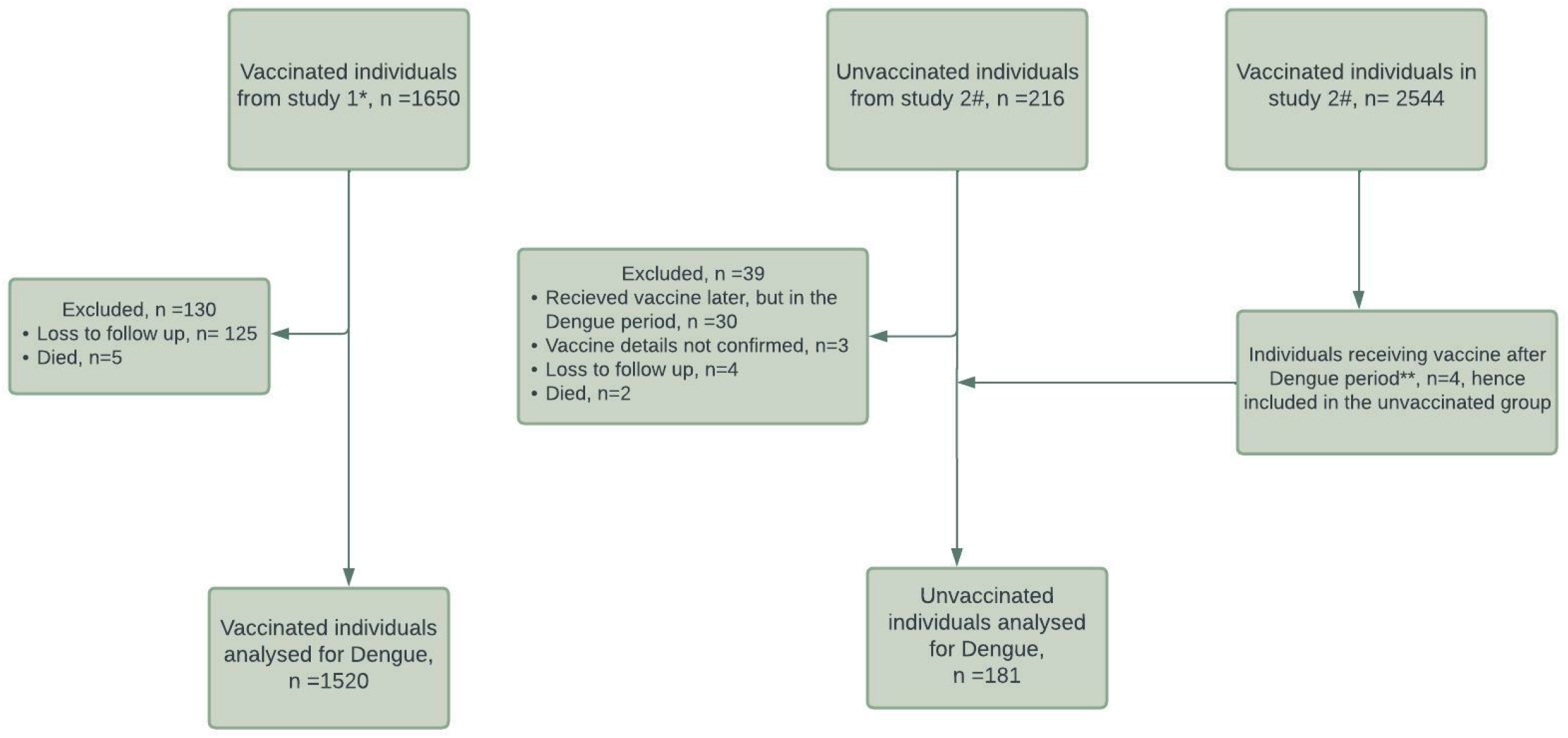
STROBE Flowchart of enrolment of study participants. [*Reference no.5, #Reference no.7, **Dengue epidemic period is considered to be from 1 July 2021 to 30 September 2021]

### 3.1. Estimates of Dengue and its severity

**Table 1a** shows the distribution of baseline characteristics and occurrence of Dengue with respect to demographics, co-morbidities, history of COVID-19 and COVID-19 vaccination status. Out of total 1701 participants, symptomatic Dengue occurred in 133 (7.8%). Among Dengue cases, ‘severe’ Dengue occurred in 42 (31.6%). A significant association for the occurrence of Dengue was observed with age and COVID-19 of the first wave (i.e., 2020). Symptomatic Dengue occurred commonly in individuals with a history of COVID-19 in the first wave and in young individuals <40 years of age. No association existed however with COVID-19 of the second wave or vaccination status. **Table 1b** shows the association of severity of Dengue with the same variables. Severity of Dengue did not share any significant association with any of the variables.

### 3.2. Risk factors of Dengue occurrence and severity

**Table 2a** shows the risk factors of occurrence of Dengue. COVID-19 in the year 2020 was an independent risk factor of symptomatic Dengue (aOR=2, p=0.002). No risk of development of Dengue was noticed with COVID-19 of the year 2021 i.e. second wave COVID-19 or with COVID-19 vaccination. Severity of Dengue was not influenced by either COVID-19 or vaccination status as shown in **Table 2b**.

### 3.3. Risk of Dengue based on timing of vaccine with respect to COVID-19

**Figures 2a and 2b** reflect the occurrence and severity of Dengue when participants were classified based on their prior history of COVID-19, vaccination status and timing of the vaccine with respect to COVID-19 of the year 2020. With statistical significance (P=0.02), highest occurrence (13.3%) of Dengue was noticed in individuals of the VAC group. The rates were low in CAV (8%), CNV (7.9%) and VNC (6.8%). The NVNC group had the lowest rates of Dengue (4.3%). No statistically significant association was observed between these groups and Dengue severity (P=0.26). 25 participants of this CAV group subsequently received their 2^nd^ dose of vaccine after COVID-19 of the year 2021 and hence were moved to VAC group of Categorization B (sensitivity analysis). Similar patterns of incidence of Dengue were observed in individuals classified as per Categorization B (**Figure 2a**).

**Figure 2a.**
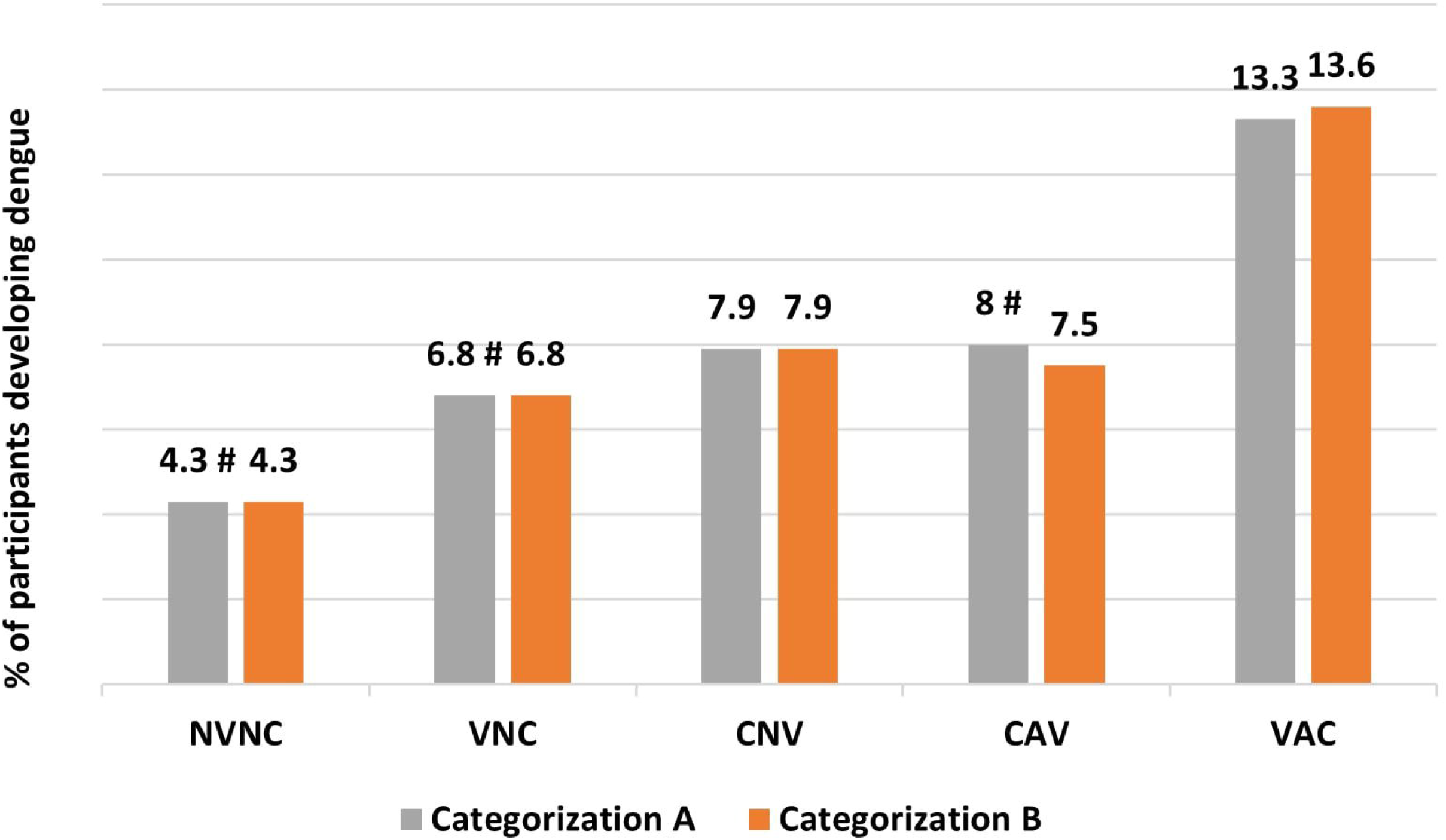
Occurrence rate of dengue in study participants categorized based on COVID-19 illness at any time (before dengue), vaccination status and timing of COVID-19 vaccine with respect to COVID-19 illness. [#P-value <0.05 compared to VAC; Abbreviations-CAV, CovidAfterVaccine, CNV, CovidNoVaccine, NVNC, NoVaccineNoCovid, VAC, VaccineAfterCovid, VNC, VaccineNoCovid]

**Figure 2b.**
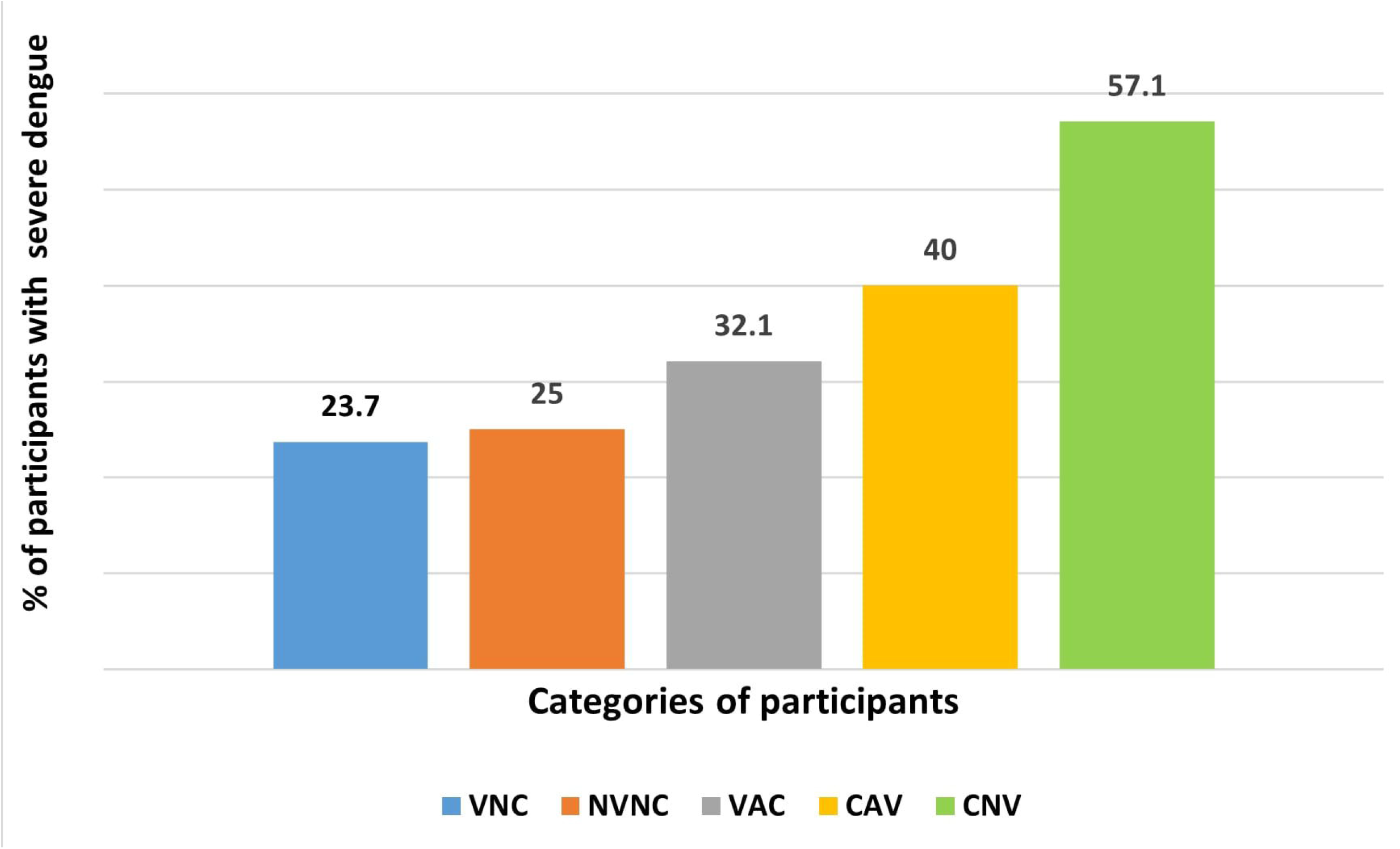
Severity of dengue in study participants categorized based on COVID-19 illness at any time (before dengue), vaccination status and timing of COVID-19 vaccine with respect to COVID-19 illness. [Abbreviations-CAV, CovidAfterVaccine, CNV, CovidNoVaccine, NVNC, NoVaccineNoCovid, VAC, VaccineAfterCovid, VNC, VaccineNoCovid]

**Table 3** shows the regression analysis results of occurrence of symptomatic Dengue. Compared to individuals with no history of COVID-19 and no history of COVID-19 vaccination (NVNC group), a 3.5 times higher odds of Dengue occurrence was observed in individuals receiving COVID-19 vaccine after recovery from prior COVID-19 of the year 2020 (VAC group) (p=0.023). In addition, the VAC group had 1.98 times and 1.74 times higher odds of Dengue compared respectively to individuals of the VNC group and those of the CAV group. Results were corroborated in another analysis as per Categorization B (**Table 3b**). A 3.6, 2.04- and 1.92-times higher odds of Dengue was observed in the VAC group compared respectively to the NVNC, VNC and CAV groups.

## 4. Discussion

Dengue continues to remain endemic in more than 100 countries with around 70 percent of the disease burden from Asia (8). In India, the disease burden has seen an unprecedented rise in recent times posing substantial challenges to the health infrastructure (4). The Dengue virus belongs to the Flavivirus genus and has four serotypes (DEN1, DEN2, DEN3, DEN4), primarily caused by the bite of infected *Aedes aegypti* mosquitoes. DEN2 is the most prevalent serotype in India (9) and though recovery from infection provides immunity against a particular serotype, secondary infection by other serotypes continues to remain a challenge, particularly in hyperendemic regions. As per CDC, one out of four infected cases is symptomatic, symptoms being governed by immune responsiveness of the host and level of viremia.

The institute where the present study was conducted faced the brunt of the 2^nd^ wave of COVID-19 during the months of March-May 2021 and Dengue during the period of July-September 2021. The present study was hence planned to explore a possible relationship between COVID-19 and Dengue. The occurrence of Dengue was two times more common in individuals with a history of COVID-19 in the first wave of the pandemic i.e. in the year 2020. There was a trend towards a higher risk of occurrence of Dengue in individuals less than 40 years of age. Interestingly, the occurrence of Dengue was not determined by COVID-19 of the second wave, nor was it affected by the vaccination status of participants. However, a significant association was observed when apart from vaccination status and history of COVID-19,individuals were categorized depending upon their cumulative COVID-19 and vaccination status and timing of vaccination with respect to COVID-19. Individuals who received the vaccine after recovery from COVID-19 had a 3.5 times higher odds of developing dengue when compared to individuals with no history of COVID-19 illness and vaccination. This same group was at higher risk of developing Dengue compared both to vaccinated individuals with no history of COVID-19 and individuals developing COVID-19 after receipt of vaccine.

The underlying mechanisms via which COVID-19 modulates the occurrence and severity of Dengue have not been fully elucidated. Mechanistic understanding of chronic immunological perturbations by SARS-CoV-2 will lend more insights on how SARS-CoV-2 may potentially enhance the vulnerability of an individual to symptomatic Dengue. Recently, a decline in CD8+ T cells and CD4+ T cells and an increase in CD16+ monocytes has been observed in COVID-19 convalescent patients even after 6-8 months of recovery from the infection (10). Previously, a low frequency of IFN-γ and IL-2 producing T cells has been linked with subclinical forms of Dengue while a high frequency of corresponding cells conferred protection from symptomatic Dengue (11). Dengue specific CD4+ and CD8+ T cells curtail the infection by the production of IFN-γ, TNF-α and IL-2 as well as by direct cytotoxicity or lysis of infected cells (12). Accordingly, the persistent lymphopenia observed in post COVID-19 state may result in uncontrolled replication of Dengue virus inside host cells. The higher rates of symptomatic Dengue in individuals with a history of COVID-19 and specifically in individuals receiving COVID-19 vaccine after natural infection, might also be explained by the phenomenon of antibody dependent enhancement (ADE) of infection. The ADE in Dengue occurs when a person after recovering from a primary infection by one serotype suffers from a secondary heterotypic infection with another serotype. The cross-reactive antibodies produced by primary infection fail to neutralize the secondary infection and instead form complexes with the new strain. The resulting interaction of the virus with Fcγ-receptors of monocytes promotes viral entry into host cells and facilitates viral replication. The level of viremia decides the clinical manifestations of Dengue (13). Classically, ADE triggers severe forms of Dengue such as Dengue haemorrhagic fever (DHF) and Dengue shock syndrome (DSS) by producing an aberrant ‘cytokine storm’. Theoretically, however, ADE decides the level of viremia and hence can also influence the patterns of clinical presentation of Dengue-whether subclinical or symptomatic. Worth investigating in continuation is whether the anti-S or anti-RBD antibodies produced in response to natural SARS-CoV-2 infection or vaccination cross react with any Dengue virus epitope which may facilitate the disease course of Dengue. Though DENV serotyping was not done in the study participants, ADE has also been projected to produce cyclical and chaotic epidemics with enhanced strains in various mathematical models on Dengue dynamics (14).

The concept of cross-reactive immune response between different viruses gains support from reports of outbreaks of Zika virus in Dengue endemic American regions. Antibodies against structural proteins of DENV cross-react with structural epitopes of the Zika virus. The subsequent interaction has been shown to produce minimal to enhancing effect on Zika infection and pathogenesis (15). Antibodies against Zika virus also show cross reactivity with DENV and enhanced Dengue occurrence and severity has been noticed in Dengue prone animal models upon administration of Zika specific antibodies (16). The cellular response mounted against DENV or Zika, on the other hand, is mutually cross protective. The predominant humoral response generated by DENGVAXIA, a yellow fever-based Dengue vaccine against the structural protein of DENV, has been viewed as a reason for a non-satisfactory protection against Dengue. Accordingly, vaccines based on non-structural proteins of DENV can rather impart a stronger protection against Dengue and possibly against Zika. Vaccination platforms employed against COVID-19 are primarily based on the mounting of humoral response against the structural spike protein of SARS-CoV-2. A rather high vulnerability of vaccinated individuals towards the original strain as well as variants of SARS-CoV-2 has been witnessed in the early months of post vaccination period (7,17,18). In this line, a vaccine based also on non-structural proteins of SARS-CoV-2 with generation of protective T cell response is a requisite in the fight against SARS-CoV-2, its variants and possibly against other viruses with similarity in epitopes.

The severity of dengue was not affected by COVID-19 or COVID-19 vaccines. Though severe forms of Dengue were numerically higher in unvaccinated individuals who had a history of COVID-19 before Dengue, relevant conclusions cannot be generated because of the small number of unvaccinated people overall and meagre representation of severe Dengue in the study. Further, severity was assessed in terms of intravenous fluid requirement or need of hospitalization. Two study participants required platelet transfusion. Among these, one had developed COVID-19 in the second wave of pandemic after being fully vaccinated and later developed severe Dengue with nadir platelet count of 10000/µL. The other had a history of COVID-19 during the second wave but was unvaccinated till the date of collection of data and developed severe Dengue with platelet count as low as 8000/µL. Mucosal bleeding in the form of epistaxis with moderate thrombocytopenia occurred in one participant who was fully vaccinated but had no history of COVID-19.

## 5. Limitations

A major drawback of the present study is the small sample size of the unvaccinated arm. Larger population level studies with a sufficient number of unvaccinated people are needed to understand the relationship between COVID-19, the COVID-19 vaccines and Dengue. Serotyping of DENV and prior history of Dengue infection were not enquired. These factors are known to influence the severity of Dengue. Seropositivity to SARS-CoV-2 or antibodies against the Spike protein of SARS-CoV-2 were not measured. This can be incorporated in the future to explore the concept of antibody dependent enhancement of DENV in the post COVID-19 era. This being a telephonic survey, detailed clinical features and blood investigations could not be performed. The severity of Dengue was assessed in terms of FDA definition of AEFI and hence specific domains of severity such as organ dysfunction or respiratory distress as highlighted in the WHO definition of severe Dengue might have been missed. Hospitalization and the need for intravenous fluids due to hypotension was enquired from all participants, along with the lowest documented platelet count or event of bleed. In addition, the study staff recorded any additional clinical feature informed by the participants, thus minimizing our chances of missing out on warning signs of Dengue. The vaccinated participants had received COVISHIELD, an Adeno virus-based vaccine. The vaccination related results of the present study hence cannot be extrapolated to other vaccines. Also, the majority of participants were health care workers and representation of co-morbidities was meagre. Individuals with co-morbidities such as cardiovascular conditions and diabetes are a higher risk of severe forms of Dengue and hence should be studied to better understand the determinants of Dengue severity. Laboratory diagnosis of Dengue was made based on the serum positivity of the NS1 antigen or IgM antibodies against DENV. Association between COVID-19 and Dengue was determined for Dengue cases diagnosed by either of the methods and subgroup association based only on NS1 or IgM was not performed. Because of the small number of cases with severe forms of COVID-19 and severe forms of Dengue, the effect of severity of COVID-19 on severity of Dengue could not be evaluated. Again, this being an exploratory analysis, Dengue statistics were not the primary outcomes of the original two studies and hence the current study might not be powered enough to comment upon the Dengue estimates.

## 6. Conclusion

COVID-19 enhanced the risk of development of Dengue by two-fold. The risk was enhanced further in individuals receiving COVID-19 vaccine after recovering from COVID-19. The possible mechanisms of immunomodulation including but not limited to T-cell suppression and antibody dependent enhancement of Dengue by SARS-CoV-2 or anti-Spike antibodies should be explored. Long term effects of COVID-19 or COVID-19 vaccines on disease course of other viral illnesses should be investigated. Larger population level studies with better representation of Dengue spectrum and co-morbidities, and adequate enrolment of unvaccinated people are needed to understand the patterns of modulation of Dengue by COVID-19, COVID-19 vaccination, and timing of vaccination with respect to COVID-19.

## Data Availability

All data produced in the present study are available upon reasonable request to the authors, as per institutional and national legal norms and procedures.

## Acknowledgements

The authors UK and SSC thank the Institutions of Eminence Scheme in the Banaras Hindu University for research support. SSC thanks the Indian Council of Medical Research for research support. UK thanks Dr Vaibhav Jaisawal and Dr Kalika Maheshwari for logistic support and Mr Gaurav Kumar Gupta for project assistance.

## Ethical Statement

The study was conducted after permission from the Institute Ethical Committee of the Institute of Medical Sciences, Banaras Hindu University. Written informed consent was taken from each participant. No human experimentation was performed. All procedures were performed as per the Declaration of Helsinki and its subsequent modifications.

