## Supplementary material for "Did COVID-19 or COVID-19 vaccines influence the patterns of Dengue in 2021: An exploratory analysis of two observational studies from North India": Table 1

| 1a | | | | 1b |  | |
| --- | --- | --- | --- | --- | --- | --- |
|  | **N=1701** | **Dengue cases, n (%) *** | **P-value** | **N= 133** | **Severe dengue,**  **n (%) ^#^** | **P-value** |
| Age (years)  < 40  ≥ 40 | 1104  597 | 97 (8.8)  36 (6) | **0.04** | 97  36 | 32 (33)  10 (27.8) | 0.56 |
| Sex  Female  Male | 563  1138 | 44 (7.8)  89 (7.8) | 0.99 | 44  89 | 17 (38.6)  25 (28.1) | 0.22 |
| BMI^##^  <25  ≥25 | 948  752 | 77 (8.1)  56 (7.4) | 0.60 | 77  56 | 26 (33.8)  16 (28.6) | 0.52 |
| COVID-19 in year 2020  No  Yes | 1470  231 | 102 (6.9)  31 (13.4) | **0.001** | 102  31 | 32 (31.4)  10 (32.3) | 0.93 |
| COVID-19 in year 2021  No  Yes | 1122  579 | 82 (7.3)  51 (8.8) | 0.27 | 82  51 | 23 (28)  19 (37.3) | 0.27 |
| Diabetes mellitus  No  Yes | 1561  140 | 123 (7.9)  10 (7.1) | 0.83 | 123  10 | 37 (30)  5 (50) | 0.28 |
| Hypertension  No  Yes | 1525  176 | 119 (7.8(  14 (8.1) | 0.84 | 119  14 | 39 (32.8)  3 (21.4) | 0.547 |
| Heart Disease  No  Yes | 1674  27 | 130 (7.8)  3 (11.1) | 0.46 | 130  3 | 41 (31.5)  1 (33.5) | 1.8 |
| Lung Disease  No  Yes | 1653  48 | 128 (7.7)  5 (10.4) | 0.42 | 128  5 | 42 (32.8)  0 | 0.18 |
| Hypothyroidism  No  Yes | 1637  64 | 128 (7.8)  5 | 0.99 | 128  5 | 41 (32)  1 (20) | 1.0 |
| Vaccination status  Vaccinated  Unvaccinated | 1520  181 | 122 (8)  11 (6.1) | 0.36 | 122  11 | 37 (30.3)  5 (45.5) | 0.32 |

**Table 1. Association of occurrence of dengue (Table 1 a) and severity of dengue (Table 1b) with demographics, co-morbidities, COVID-19, and COVID-19 vaccination.**

*Percentage calculated out of total participants analyzed (N=1701) ^#^Percentage calculated out of dengue cases (N=133). ^##^Not known for one participant.
