## Supplementary material for "Did COVID-19 or COVID-19 vaccines influence the patterns of Dengue in 2021: An exploratory analysis of two observational studies from North India": Table 2

| 2a Risk factors of occurrence of Dengue | | |  | 2b Risk factors of severity of Dengue | | |
| --- | --- | --- | --- | --- | --- | --- |
| n =1701 | **aOR (CI)** | **P-value** |  | **n = 133** | **aOR (CI)** | **P-value** |
| Age (years)  <40  ≥40 (Reference) | 1.44 (0.96–2.16) | 0.07 |  | **Any COVID-19**  Yes No (Reference) | 2.0  (0.93- 4.17) | 0.08 |
| COVID-19 in year 2020  Yes  No (Reference) | **2** (1.30–3.09) | **0.002** |  |  |  |  |
| COVID-19 in year 2021  Yes  No (Reference) | 1.2 (0.84–1.75) | 0.31 |  | **Vaccination status**  Unvaccinated  Vaccinated (Reference) | 1.8 (0.5–6.3) | 0.36 |
| Vaccination status  Unvaccinated  Vaccinated (Reference) | 0.69 (0.36–1.32) | 0.27 |  |  |  |  |

**Table 2. Tentative risk factors of occurrence (Table 2a) and severity (Table 2b) of Dengue in adjusted analysis (logistic regression).**

**aOR: adjusted Odds ratio, CI: confidence interval**
