## Supplementary material for "Did COVID-19 or COVID-19 vaccines influence the patterns of Dengue in 2021: An exploratory analysis of two observational studies from North India": Table 3

|  | 3a* |  | 3b^#^ |  |
| --- | --- | --- | --- | --- |
| Risk factors | **aOR (CI)** | **P-value** | **aOR (CI)** | **P-value** |
| Age (years)  <40  ≥40 (Reference) | 1.46 (0.97-2.2) | 0.065 | 1.46 (0.98-2.2) | 0.06 |
| Categories of participants | | |  |  |
| NVNC (Reference)  VAC  CNV  VNC  CAV | **3.5** (1.2-10.2)  1.85 (0.52-6.5)  1.7 (0.62-4.9)  2 (0.69-5.8) | **0.023**  0.34  0.29  0.19 | **3.6** (1.2-10.5)  1.85 (0.52-6.5)  1.75 (0.62-4.96)  1.87 (0.64-5.4) | **0.019**  0.34  0.29  0.25 |
| VNC (Reference)  VAC | **1.98** (1.23-3.20) | **0.005** | **2.04** (1.3-3.2) | **0.002** |
| CAV (Reference)  VAC | **1.74** (1.02-2.9) | **0.04** | **1.92** (1.14-3.23) | **0.015** |
| CNV (Reference)  VAC | 1.8 (0.79-4.5) | 0.15 | 1.94 (0.82-4.58) | 0.13 |

**Table 3. Risk factors of occurrence of dengue depending on categorization of participants based on COVID-19 at any time, vaccination status and timing of vaccine with respect to COVID-19 episode.**

***Table 3a, VAC includes individuals with history of receiving vaccine in 2021 after COVID-19 of the year 2020**

**^#^Table 3b, VAC includes individuals with history of receiving COVID-19 vaccine in 2021 after COVID-19 of the year 2020 as well as individuals receiving 2^nd^ dose of COVID-19 vaccine after COVID-19 of the year 2021, but before development of Dengue**

a OR: adjusted Odds ratio, CAV: CovidAfterVaccine, CI: confidence interval, CNV: CovidNoVaccine, NVNC: NoVaccineNoCovid, VAC: VaccineAfterCovid, VNC: VaccineNoCovid
